# Fluvoxamine for Outpatient COVID-19 to Prevent Hospitalization: A Systematic Review and Meta-Analysis

**DOI:** 10.1101/2021.12.17.21268008

**Authors:** Todd C. Lee, Simone Vigod, Émilie Bortolussi-Courval, Ryan Hanula, David R. Boulware, Eric J. Lenze, Angela M. Reiersen, Emily G. McDonald

## Abstract

**Importance:** Widely available and affordable options for the outpatient management of COVID-19 are needed, particularly therapies that prevent hospitalization.

**Objective:** Perform a meta-analysis of the available randomized clinical trial evidence for fluvoxamine in the outpatient management of COVID-19.

**Data Sources:** World Health Organization International Clinical Trials Registry Platform and ClinicalTrials.gov.

**Study Selection:** Completed outpatient trials with available results which compared fluvoxamine to placebo.

**Data Extraction and Synthesis:** We followed the PRISMA 2020 guidelines. We extracted study details in terms of inclusion criteria, trial demographics and the pre-specified outcome of all-cause hospitalization. Risk of bias was assessed by the Cochrane Risk of Bias 2 tool. We conducted a frequentist random effects meta-analysis, as well as two sensitivity analyses using a Bayesian random effects meta-analysis with different estimates of prior probability: a weakly neutral prior (50% chance of efficacy with 95% confidence interval for Risk Ratio [RR] between 0.5 and 2) and a moderately optimistic prior (85% chance of efficacy). We contextualized the results by estimating the probability of any effect (RR ≤1) and moderate effect (RR ≤0.9) on reducing hospitalization.

**Main Outcome(s) and Measure(s):** All cause hospitalization.

**Results:** 2196 participants were included from 3 identified trials. The risk ratios for hospitalization were 0.75 (95%CI, 0.57-0.97) for the frequentist analysis, 0.78 (95%CI 0.58-1.08) for the Bayesian weakly neutral prior, and 0.73 (95%CI, 0.53-1.01) for the Bayesian moderately optimistic prior. Depending on the scenario, the probability of any effect on hospitalization ranged from 94.1% to 98.3% and a moderate effect from 81.6% to 91.1%.

**Conclusions and Relevance:** Under a variety of assumptions, fluvoxamine shows a high probability of preventing hospitalization in outpatients with COVID-19. While ongoing randomized trials are important to evaluate alternative doses, explore the effectiveness in vaccinated patients, and provide further refinement to these estimates, fluvoxamine could be recommended as a treatment option, particularly in resource-limited settings or persons without access to SARS-CoV-2 monoclonal antibody therapy or direct antivirals.

**Key Points:** *Question:* Does early administration of fluvoxamine prevent hospitalization in symptomatic adult outpatients with confirmed COVID-19?

*Findings:* In this meta-analysis with Bayesian sensitivity analyses that accounted for varying prior probabilities, there was a high probability (94.1% to 98.3%) that fluvoxamine reduces hospitalization with frequentist risk ratio of 0.75 (95%CI 0.57-0.97).

*Meaning:* Fluvoxamine is a widely available and inexpensive option that prevents hospitalization in patients with early COVID-19 based on randomized controlled trial evidence to date.

## Introduction

Finding effective outpatient therapies for COVID-19 has been a major research undertaking since the beginning of the pandemic. While therapies such as direct antivirals and engineered monoclonal antibodies represent the current state-of-the art, there are currently challenges with availability, access, administration, and affordability in most areas of the world. Drug repurposing, or the use of existing available and affordable medications for the treatment of COVID-19, is an area of substantial ongoing research interest. The first medication to gain international interest for repurposing was hydroxychloroquine; however, it was ultimately shown to be ineffective in randomized controlled trials^1^. A variety of other candidate molecules have been the subject of randomized controlled trials with varying success.^2^

One such candidate is fluvoxamine, a selective serotonin reuptake inhibitor (SSRI) that is also a potent activator of the sigma-1 receptor which decreases inflammation via reducing endoplasmic reticulum stress.^3^ In a murine sepsis model, fluvoxamine administration reduced mortality predominantly through this mechanism.^4^ On this preclinical basis, the phase 2 STOP COVID 1 trial (NCT04342663) was conducted.^5^ This 152 patient double-blind, randomized placebo-control trial found fluvoxamine effective at preventing progression to severe COVID-19 defined as: hypoxemia with dyspnea and/or hospital admission. Shortly thereafter, a benefit in preventing hospitalization/death was also reported in an uncontrolled 113 person prospective cohort.^6^

Two larger phase 3 trials have been subsequently completed: STOP COVID2 in the US and Canada (NCT04668950) and the TOGETHER trial in Brazil (NCT04727424).^7^ Both trials were presented to the U.S. National Institutes of Health in August 2021. STOP COVID 2 was stopped for futility in May 2021 after an interim analysis found that the low event rate seen in the trial was associated with a <10% conditional probability of demonstrating efficacy within an attainable sample size based on recruitment rate.^8^ TOGETHER, which had a higher primary outcome event rate, was stopped after demonstrating clinical benefit.^7^ We conducted a systematic review and meta-analysis of outpatient fluvoxamine trials to contextualize the evidence with respect to hospitalization to inform clinical decision making, policy, and guidelines.

## Methods

This systematic review and meta-analysis is reported according to PRISMA 2020.^9^

### Search Strategy, Study selection, and Data Extraction

On November 12, 2021, we searched the World Health Organization International Clinical Trials Registry Platform and ClinicalTrials.gov for all registered clinical trials of fluvoxamine for the treatment of patients with COVID-19. Two independent reviewers screened results for eligibility which included: completed studies of outpatients comparing fluvoxamine to placebo or standard of care. Studies with published or publicly presented data (with the authors’ permission) were selected for inclusion.

For included studies we summarized study inclusion criteria and patient demographics. We chose all-cause hospitalization as the primary outcome of interest given the implications for healthcare resource utilization. Two reviewers extracted hospitalization outcome data in each treatment group and these numbers were subsequently verified with the study principal investigators by email. For TOGETHER, the published primary outcome included hospitalization or emergency room observation lasting ≥6 hours. To arrive at a more homogenous outcome, we contacted the TOGETHER trial authors and obtained outcome data on emergency room visits lasting >24 hours and used this as a more representative proxy for hospital admission than ER visit alone. For STOP COVID 2, we obtained demographic and outcomes data directly from the principal investigators (EJL and AMR) based on their presentation to the National Institutes of Health. We only included intention-to-treat analyses even if per protocol analyses suggested larger effect sizes were possible.

### Assessment of Bias

Two independent reviewers assessed each study for bias using the Cochrane risk-of-bias 2 tool for randomized trials.

### Meta-Analysis

We first conducted a frequentist DerSimonian–Laird random effects meta-analysis using STATA version 17 (StataCorp, USA) on the risk ratio scale. With low heterogeneity this would give identical results to a fixed effects model with inverse variance weights. Then, to be more conservative, we conducted two sensitivity analyses with a Bayesian random effects meta-analysis on the log risk ratio scale using *bayesmeta* in R version 4.1.1.^10^ We selected priors based on two estimates of how promising the pre-existing data were. Scenario 1 used a weakly informative neutral prior, given that most treatment effects in medicine fall within this range and because of the STOP COVID 2 results (with a mean mu 0 and a standard deviation 0.355 this corresponds to a 50% chance of efficacy with 95% probability that the risk ratio (RR) would be between 0.5 and 2).^11^ Scenario 2 used a moderately optimistic prior given the positive results of STOP COVID 1 and the prospective cohort (with a mean mu -0.41 and a standard deviation 0.4 this corresponds to a 85% chance the RR would be ≤1).^11^ In both scenarios, the prior for the heterogeneity parameter (tau) used a half-Cauchy distribution with scale 0.10 which is the average heterogeneity for meta-analyses of trials using hospitalization outcomes.^12^ Final results were exponentiated to the risk ratio scale for presentation.

The forest plot for the random effects meta-analysis and graphs of the prior and posterior probability distributions based on the pooled RRs were created with STATA version 17 (StataCorp, USA). Probabilities of any effect (RR<1) and a moderate effect (RR ≤0.9) were calculated by integrating the area under the posterior probability density curves.^13^ Given the low cost of fluvoxamine and decades of established safety, we decided in advance that a moderate effect would correlate to an absolute risk reduction between 0.5% and 1% assuming a 5-10% baseline risk of hospitalization, as observed in the control groups of several outpatient clinical trials. For context, this corresponds to a Number Needed to Treat (NNT) of 100-200.

### Certainty Assessment

Two independent reviewers assessed the certainty of evidence for hospitalization using GRADE methodology (Grading of Recommendations, Assessment, Development and Evaluations)^14^.

## Results

The initial search yielded 19 candidate randomized controlled trials and 10 were retained following removal of duplicates **(Supplemental Figure 1** and **Supplemental Table 1)**. Seven were then excluded for the following reasons: 4 studies because they were still recruiting (NCT04510194, NCT04718480/EUCTR2020-002299-11-HU, NCT04885530, and NCT05087381), 1 study because it recruited only inpatients (IRCT20131115015405N4), 1 study because it was suspended without results (NCT04711863), and 1 was excluded because it had not yet started recruitment (TCTR20210615002).

### Included studies

The remaining 3 trials (STOP COVID 1 (n=152)^5^, STOP COVID 2 (n=547), and TOGETHER (n=1497)^7^) included a total of 2196 analyzed patients (**Table 1**). All 3 were placebo-controlled randomized controlled trials that recruited unvaccinated, symptomatic adults with microbiologically confirmed SARS-CoV-2 infection who were within 6-7 days of infection and not requiring oxygen. Whereas STOP COVID 1 took all patients, STOP COVID 2 and TOGETHER enriched their population for at least one at-risk feature for deterioration. Overall, the median age of subjects was between 46 and 50, 55-72% of participants were female, 44-56% were obese. Most patients in the STOP COVID trials self-reported as Caucasian; in TOGETHER, 96% self-reported as mixed race. The risk of bias was considered low for all trials by both reviewers.

**Table 1.** Fluvoxamine COVID-19 Trial Details.

| Study | Location | Original Outcome | Inclusion Criteria | Demographics | Fluvoxamine target dose | Duration |
| --- | --- | --- | --- | --- | --- | --- |
| Stop Covid 1 | USA | Clinical deterioration: Hospitalization or new hypoxemia within 15 days | Age $\geq 18$<br>unvaccinated Positive test with:<br>$\leq 7$ days symptoms | Median age 46; 72% female;<br>70% white; 56% BMI$\geq 30$;<br>20% hypertension; 11% diabetes<br>Median 4 days of symptoms | 100mg PO TID | 15 days |
| Stop Covid 2 | USA and Canada | Clinical deterioration: Hospitalization or new hypoxemia within 15 days | Age $\geq 30$<br>unvaccinated<br>Positive test with:<br>$\leq 6$ days symptoms<br>Criterion for high risk | Median age 47; 62% female;<br>73% white; 44% BMI$\geq 30$;<br>21% hypertension; 9% diabetes<br>Median 5 days of symptoms | 100 md PO BID | 15 days |
| Together | Brazil | ER visit $\geq 6$ hours or hospitalization within 28 days | Age $\geq 18$<br>unvaccinated<br>Positive test with:<br>$\leq 7$ days symptoms<br>Criterion for high risk | Median age 50; 55% female;<br>96% mixed race; 51% BMI$\geq 30$;<br>13% hypertension; 16% diabetes<br>Mean 3.8 days of symptoms* | 100mg PO BID | 10 days |
| ER=Emergency Room; *Median not provided; missing data on ~23% |  |  |  |  |  |  |

### Meta-analysis

In the frequentist meta-analysis, the pooled RR in favor of fluvoxamine was 0.75 (95%CI 0.57-0.97; I^2^ 0.2%) (**Figure 1**). Correspondingly, there was a 98.3% probability that fluvoxamine prevents hospitalization and a 91.1% probability of at least a moderate effect. In the Bayesian sensitivity analysis, the RR in favor of fluvoxamine was 0.78 (95%CI 0.58-1.08) for the weakly neutral prior, and 0.73 (95%CI 0.53-1.01) for the moderately optimistic prior **(Table 2)**. The probability of any effect ranged from 94.1% to 98.3%, and of moderate effect from 81.6% to 91.1% (**Supplemental Figure 2**).

**Table 2.**
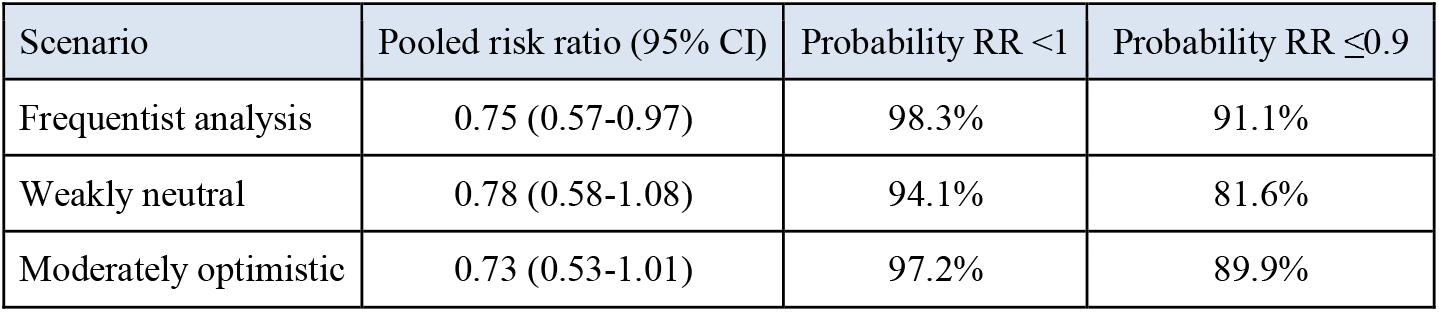
Meta-analysis results.

| Scenario | Pooled risk ratio (95% CI) | Probability RR <1 | Probability RR ≤0.9 |
| --- | --- | --- | --- |
| Frequentist analysis | 0.75 (0.57-0.97) | 98.3% | 91.1% |
| Weakly neutral | 0.78 (0.58-1.08) | 94.1% | 81.6% |
| Moderately optimistic | 0.73 (0.53-1.01) | 97.2% | 89.9% |

**Figure 1.**
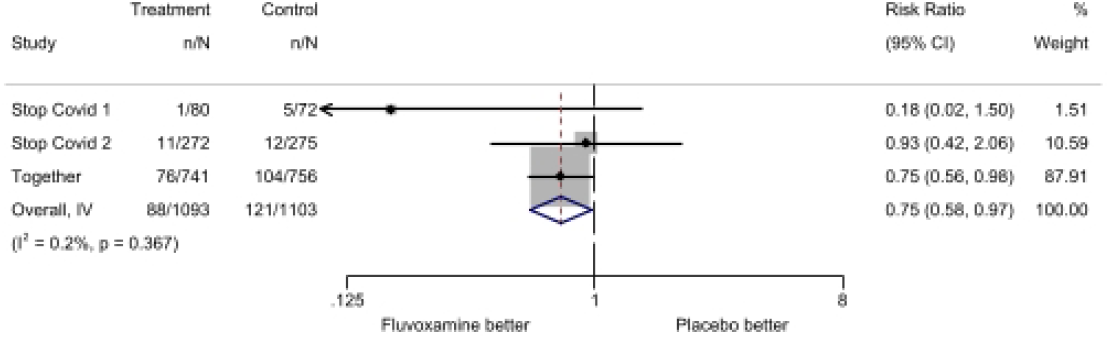
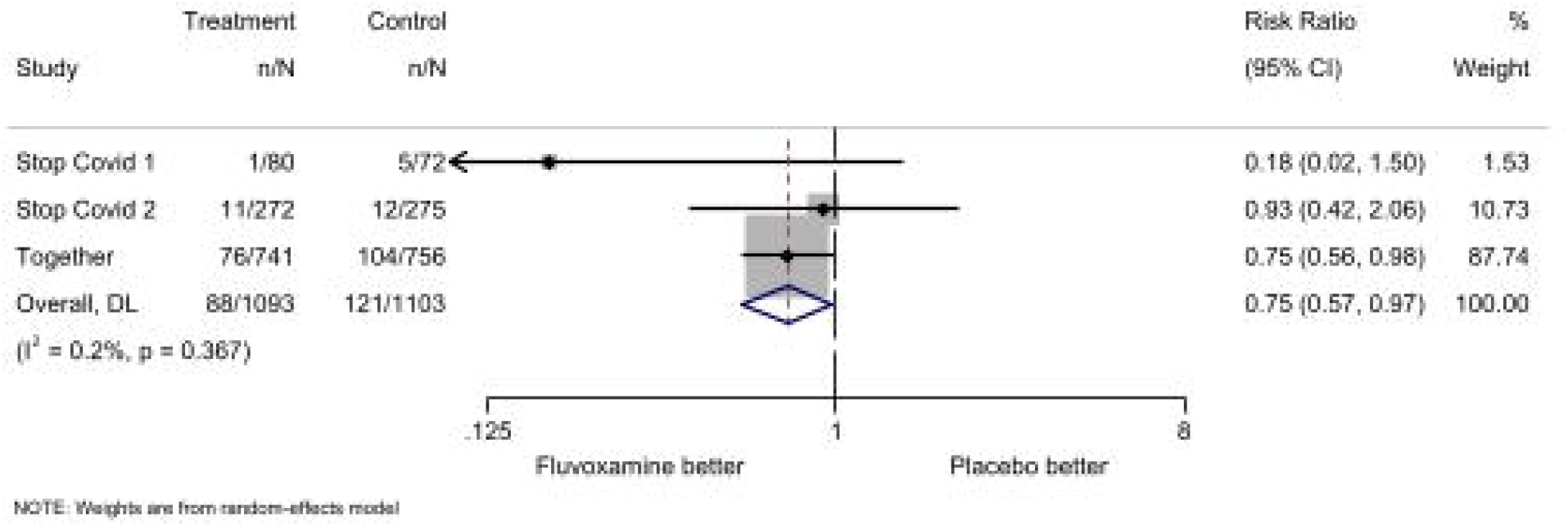
Frequentist Random Effects Meta-Analysis.

### GRADE Certainty of Evidence

We believe the certainty of the evidence is moderate. Although all three trials were placebo-controlled randomized trials, there is some inconsistency in the findings given STOP COVID 2 was terminated for futility. Although there is some risk of bias in the TOGETHER trial, we attempted to address this by limiting the primary outcome to emergency room visits that were 24 hours or longer, thus removing some of the concern for bias that could have been introduced through the inclusion of shorter visits (6-24 hours in length). A moderate strength recommendation in favor of fluvoxamine could be considered in certain clinical scenarios detailed in the discussion. According to the GRADE methodology, such a recommendation is reasonable when shared decision making is expected. Weaker recommendations usually apply when risks start to approach the benefits, or when significant resources are required for the intervention, neither of which is the case for fluvoxamine.

## Discussion

Based on all currently available clinical trial data, it is very likely that fluvoxamine has at least a moderate effect on preventing COVID-19 hospitalization under a variety of assumptions. By comparison, outpatient trials with hydroxychloroquine^1^ and ivermectin^15^ have not shown efficacy and yet these agents continue to be prescribed. Further RCT data from the ongoing identified studies like CovidOut (NCT04510194) and ACTIV-6 (NCT04885530) will help refine these estimates; however, both trials are using 50mg twice daily which, if unsuccessful, could indicate that 100mg twice daily is the minimum effective dose. Recently, based on STOP COVID 1 and the TOGETHER trial, the Infectious Diseases Society of America recommended against the use of fluvoxamine outside of clinical trial settings.^16^ Based on our analysis and coupled with worldwide accessibility, decades of safety data, and a current price of approximately $1/day,^17,18^ fluvoxamine seems reasonable for high-risk outpatients who do not have access to SARS-CoV-2 monoclonal antibodies, direct antivirals, or clinical trials. Even at a number to treat of 200 (absolute risk reduction 0.5%) the corresponding cost to prevent admission would only be $2800. Clinicians who prescribe fluvoxamine for COVID-19 should familiarize themselves with relative contraindications and drug-drug interactions, including the need to limit caffeine **(Supplemental Table 2)**.

Strengths of our analysis are the use of all available data and consistency of results by both a classic random effects meta-analysis and a Bayesian approach with multiple prior probability estimates. For all analyses, we quantified the overall probability of any effect and a moderate effect of fluvoxamine on hospitalization to help with decision making. Further, we restricted the TOGETHER trial emergency room visits to those >24 hours to address concerns^16^ about whether a six-hour visit is an accurate proxy for healthcare utilization or marker of clinical deterioration.

We utilized the intention-to-treat analysis, reflecting physician prescribing. In the TOGETHER trial, results were better in the per-protocol analysis.^7^

Limitations include variability in healthcare practices, resource availability, and circulating variants between trials, leading to differences in the baseline event rates and the associated absolute risk reduction. While we have attempted to correct for subjectivity by limiting this analysis to hospitalization, indeed hospitalization decisions may vary between geographic areas and even time points based on systemic burden. Nonetheless, all-cause hospitalization is the most common important outcome of outpatient COVID-19 trials because ICU admission or death would require studies that were prohibitively large. Additionally, all three trials excluded fully vaccinated individuals, whose rates of hospitalization are greatly reduced, and therefore any estimates of absolute effect size would likely be an overestimate in vaccinated patients. Another limitation is the inclusion of only 3 trials to date. Using a living systematic review approach will address this clinical question. This is planned in order to address this concern and to rapidly incorporate emerging evidence.

Ongoing randomized controlled trials of fluvoxamine should continue, particularly those studying lower 50mg doses (which will be better tolerated), evaluating efficacy in vaccinated individuals, or studying the related SSRI fluoxetine which is on the World Health Organization’s list of essential medications. In the meantime, fluvoxamine is an immediately available, safe, and inexpensive treatment option with a high probability of moderate efficacy. It could be recommended as a treatment option for patients without contraindication, particularly in resource-limited settings or those without access to monoclonal antibodies or direct antivirals.

## Supporting information

PRISMA Checklist

## Data Availability

All data produced in the present work are contained in the manuscript

## Acknowledgements

TCL and EGM receive research salary support from the Fonds de Recherche du Québec – Santé.

## Registration

Initially conducted as a rapid review and then changed to a full systematic review and meta-analysis, this study was not registered, and a full review protocol was not written.

## Availability of Data, Code and Other Materials

All relevant source data is contained within. Statistical code needed to reproduce the analysis is available upon written request.

## CRediT author statement

Conceptualization - TCL, EGM; Methodology - TCL, EGM; Validation - TCL; Formal Analysis - TCL; Investigation - All authors; Resources - TCL; Data Curation - TCL, EGM; Writing - Original Draft - TCL, EGM; Writing - Review and Editing - All authors; Visualization TCL, EGM

## References

1. Skipper CP, Pastick KA, Engen NW, et al. Hydroxychloroquine in Nonhospitalized Adults With Early COVID-19□: A Randomized Trial. Ann Intern Med. 2020;173(8):623–631. doi:10.7326/M20-4207

2. Ezer N, Belga S, Daneman N, et al. Inhaled and intranasal ciclesonide for the treatment of covid-19 in adult outpatients: CONTAIN phase II randomised controlled trial. BMJ. 2021;375:e068060. doi:10.1136/bmj-2021-068060

3. Sukhatme VP, Reiersen AM, Vayttaden SJ, Sukhatme VV. Fluvoxamine: A Review of Its Mechanism of Action and Its Role in COVID-19. Front Pharmacol. 2021;12:652688. doi:10.3389/fphar.2021.652688

4. Rosen DA, Seki SM, Fernández-Castañeda A, et al. Modulation of the sigma-1 receptor-IRE1 pathway is beneficial in preclinical models of inflammation and sepsis. Sci Transl Med. 2019;11(478):eaau5266. doi:10.1126/scitranslmed.aau5266

5. Lenze EJ, Mattar C, Zorumski CF, et al. Fluvoxamine vs Placebo and Clinical Deterioration in Outpatients With Symptomatic COVID-19: A Randomized Clinical Trial. JAMA. 2020;324(22):2292–2300. doi:10.1001/jama.2020.22760

6. Seftel D, Boulware DR. Prospective Cohort of Fluvoxamine for Early Treatment of Coronavirus Disease 19. Open Forum Infect Dis. 2021;8(2):ofab050. doi:10.1093/ofid/ofab050

7. Reis G, Dos Santos Moreira-Silva EA, Silva DCM, et al. Effect of early treatment with fluvoxamine on risk of emergency care and hospitalisation among patients with COVID-19: the TOGETHER randomised, platform clinical trial. Lancet Glob Health. Published online October 27, 2021:S2214-109X(21)00448-4. doi:10.1016/S2214-109X(21)00448-4

8. Lenze EJ. August 20, 2021: Fluvoxamine for Early Treatment of COVID-19: The STOP COVID Clinical Trials (Eric Lenze, MD). Rethinking Clinical Trials. Published August 24, 2021. Accessed November 1, 2021. https://rethinkingclinicaltrials.org/news/august-20-2021-fluvoxamine-for-early-treatment-of-covid-19-the-stop-covid-clinical-trials-eric-lenze-md/

9. Page MJ, McKenzie JE, Bossuyt PM, et al. The PRISMA 2020 statement: an updated guideline for reporting systematic reviews. BMJ. 2021;372:n71. doi:10.1136/bmj.n71

10. Röver C. Bayesian Random-Effects Meta-Analysis Using the bayesmeta R Package. J Stat Softw Vol 1 Issue 6 2020. Published online April 27, 2020. https://www.jstatsoft.org/v093/i06

11. Zampieri FG, Casey JD, Shankar-Hari M, Harrell FE, Harhay MO. Using Bayesian Methods to Augment the Interpretation of Critical Care Trials. An Overview of Theory and Example Reanalysis of the Alveolar Recruitment for Acute Respiratory Distress Syndrome Trial. Am J Respir Crit Care Med. 2021;203(5):543–552. doi:10.1164/rccm.202006-2381CP

12. Turner RM, Jackson D, Wei Y, Thompson SG, Higgins JPT. Predictive distributions for between-study heterogeneity and simple methods for their application in Bayesian meta-analysis. Stat Med. 2015;34(6):984–998. doi:10.1002/sim.6381

13. Lee TC, McDonald EG, Butler-Laporte G, Harrison LB, Cheng MP, Brophy JM. Remdesivir and systemic corticosteroids for the treatment of COVID-19: A Bayesian re-analysis. Int J Infect Dis. 2021;104:671–676. doi:10.1016/j.ijid.2021.01.065

14. Guyatt GH, Oxman AD, Vist GE, et al. GRADE: an emerging consensus on rating quality of evidence and strength of recommendations. BMJ. 2008;336(7650):924–926. doi:10.1136/bmj.39489.470347.AD

15. López-Medina E, López P, Hurtado IC, et al. Effect of Ivermectin on Time to Resolution of Symptoms Among Adults With Mild COVID-19: A Randomized Clinical Trial. JAMA. 2021;325(14):1426–1435. doi:10.1001/jama.2021.3071

16. Bhimraj A, Morgan R, Hirsch Shumaker A, et al. Infectious Diseases Society of America Guidelines on the Treatment and Management of Patients with COVID-19 Version 5.5.2. Published November 5, 2021. Accessed November 8, 2021. https://www.idsociety.org/practice-guideline/covid-19-guideline-treatment-and-management/

17. Régie de l’assurance maladie du Québec (RAMQ). List of medications, September 29, 2021. List of medications, September 29, 2021. Published October 1, 2021. Accessed October 27, 2021. https://www.ramq.gouv.qc.ca/en/media/12091

18. Fluvoxamine Prices, Coupons & Savings Tips. GoodRx. Accessed November 8, 2021. https://www.goodrx.com/fluvoxamine

