## Supplemental Tables and Figures for "Fluvoxamine for Outpatient COVID-19 to Prevent Hospitalization: A Systematic Review and Meta-Analysis"

**Supplemental Figure 1 – PRISMA diagram**

**Identification of studies via databases and registers**

Records removed *before screening*:

Duplicate records (n =8)

Clear database error (n=1)

Records identified from:

WHO International Clinical Trials Registry (n=11)

Clnicaltrials.gov (n =8)

**Identification**

Records screened

(n=9)

Records excluded**

Not outpatients (n = 1)

Reports not retrieved

Ongoing recruitment (n=4)

Suspended not reported (n=1)

Not Started (n=1)

Reports sought for retrieval

(n =8)

**Screening**

Reports assessed for eligibility

(n =3)

Excluded (n=0)

Studies included in review

(n =3)

Reports of included studies

(n =2)

**Included**

*From:*  Page MJ, McKenzie JE, Bossuyt PM, Boutron I, Hoffmann TC, Mulrow CD, et al. The PRISMA 2020 statement: an updated guideline for reporting systematic reviews. BMJ 2021;372:n71. doi: 10.1136/bmj.n71

**Supplemental Figure 2 - Probability Densities**

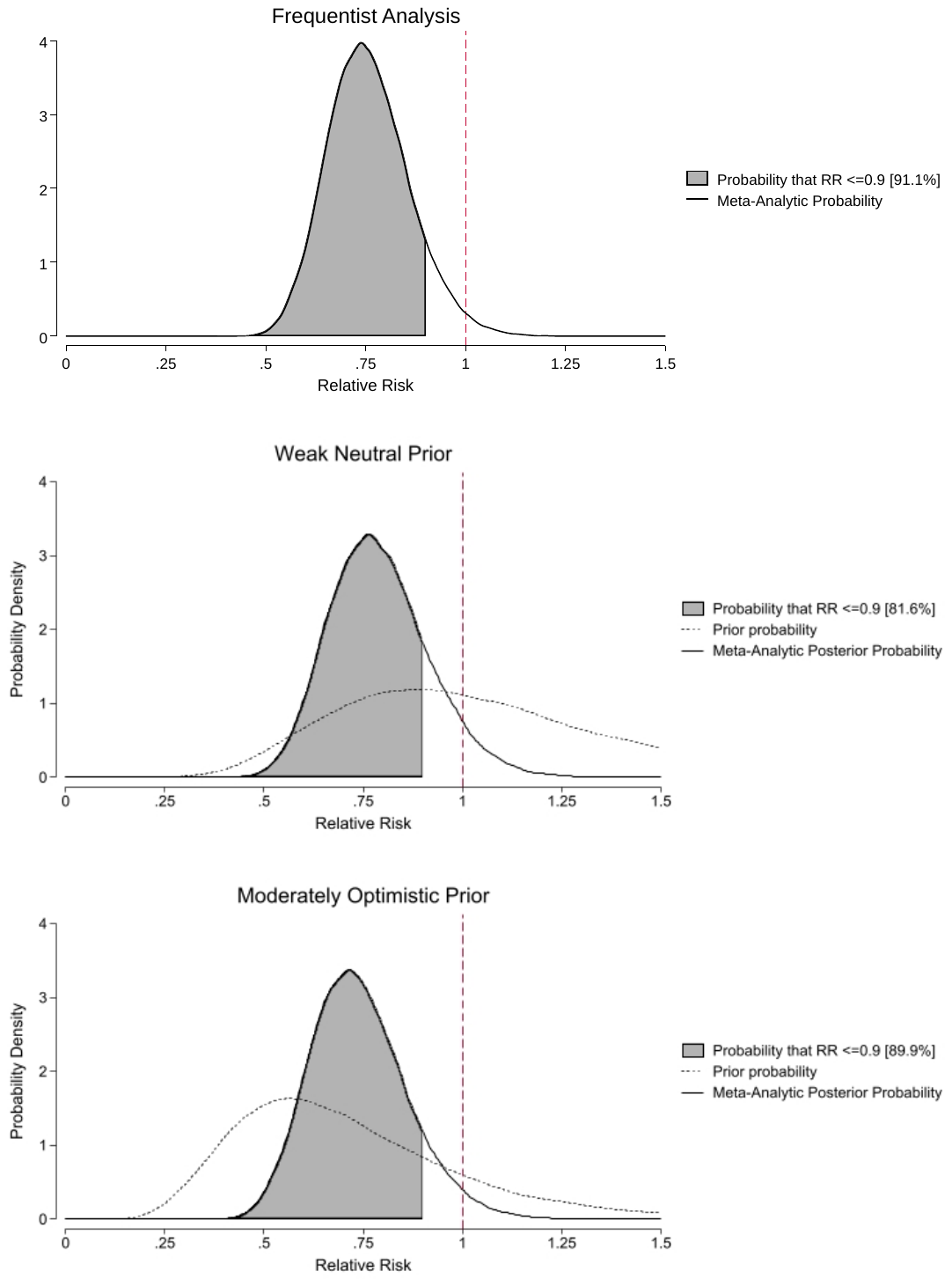

**Supplementary Table 1 –**

| Title | Registration ID(s) | Countries | Outpatient | Maximum Daily Dose and Duration | Comparator | Status | Results Available | Included in meta-analysis |
| --- | --- | --- | --- | --- | --- | --- | --- | --- |
| Effect of fluvoxamine on cytokine in COVID-19 patients | IRCT20131115015405N4 | Iran | No | 300mg x Not Specified | Standard of Care | Completed | NA | No |
| A Double-blind, Placebo-controlled Clinical Trial of Fluvoxamine for Symptomatic Individuals With COVID-19 Infection (STOP COVID) | NCT04342663 | USA | Yes | 300mg x15 days | Placebo | Completed | Yes | Yes |
| COVID-OUT: Early Outpatient Treatment for SARS-CoV-2 Infection (COVID-19) | NCT04510194 | USA | Yes | 100mg x10 days | Placebo | Recruiting | No | No |
| Fluvoxamine for Early Treatment of Covid-19 (Stop Covid 2) | NCT04668950 | USA and Canada | Yes | 200mg x15 days | Placebo | Completed | Yes | Yes |
| Fluvoxamine for Adults With Mild to Moderate COVID-19 | NCT04711863 | Republic of Korea | Yes | 200mg x10days | Placebo | Suspended | No | No |
| Fluvoxamine Administration in Moderate SARS-CoV-2 (COVID-19) Infected Patients | NCT04718480 and EUCTR2020-002299-11-HU | Hungary | Yes | 200mg x74 days | Placebo | Recruiting | No | No |
| Repurposed Approved and Under Development Therapies for Patients With Early-Onset COVID-19 and Mild Symptoms (TOGETHER) | NCT04727424 | Brazil | Yes | 200mg x10 days | Placebo | Completed | Yes | Yes |
| ACTIV-6: COVID-19 Study of Repurposed Medications | NCT04885530 | USA | Yes | 100mg x10 days | Placebo | Recruiting | No | No |
| Randomized-controlled Trial of the Effectiveness of COVID-19 Early Treatment in Community | NCT05087381 | Thailand | Yes | 150mg x14 days | Standard of Care | Recruiting | No | No |
| Effect of Combined Fluvoxamine with Favipiravir versus Favipiravir Monotherapy in Prevention of Clinical Deterioration among mild to moderate COVID-19 patients Monitoring by Telemedicine in Virtual Clinic: Open-label Randomized Controlled Trial | TCTR20210615002 | Thailand | Yes | 200mg x 10 days | Favipiravir | Not Started | NA | No |

**Supplementary Table 2: Considerations for Relative Contraindications to Fluvoxamine**

| Patient Factors | Reasoning (if not obvious) |
| --- | --- |
| Allergy to fluvoxamine |  |
| Moderate to severe depression within 6 weeks of enrollment | If the patient would need to be switched to fluvoxamine from another agent due to drug-interactions, this would ideally be done with explicit supervision |
| Previous or current diagnosis of manic depression / bipolar disorder | If the patient would need to be switched to fluvoxamine from another agent or if there would be concern that adding fluvoxamine might trigger a manic episode |
| Hepatic impairment defined as known Cirrhosis of any severity | Fluvoxamine metabolism is altered in patients with cirrhosis |
| Hospitalization for gastrointestinal or other non-traumatic bleeding within the last year | Fluvoxamine can impact platelet aggregation and these patients were excluded from the trial. This decision could be individualized. |
| Concurrent Medications |  |
| Caffeine | Fluvoxamine leads to substantial increases in caffeine levels. In the trial, we encouraged no caffeine for participants. At the very least they were told avoid more than 1 small cup of coffee’s worth of caffeine (and to stop caffeine if they felt it was “too energizing”). |
| Patients taking warfarin | Increased bleeding risk due to increased AUC of warfarin |
| Patients taking clopidogrel | Increased risk of ischemic event due to metabolism |
| Patients taking 2 or more of the following: aspirin, NSAIDS, ticlopidine, prasugrel, ticagrelor, direct oral anticoagulants | Assuming NSAIDs cannot be held. Fluvoxamine can impact platelet aggregation and these patients were excluded from the trial. This decision could be individualized. |
| Donepezil | This is a Sigma-1-receptor (S1R) agonist and we excluded patients from the trial given that fluvoxamine was being used for its S1R activity |
| Other antidepressant medications | For any patient already on a tricyclic antidepressant, SSRI, or SNRI, we evaluated whether it could be held or reduced under medical supervision during the time they were prescribed fluvoxamine. If the patient was taking a low dose of another medication (e.g., citalopram 10mg) and there was low risk of serotonin syndrome, concurrent use was allowed. |
| Use within 14 days of an MAO inhibitor [e.g., Isocarboxazid (Marplan), Phenelzine (Nardil), Selegiline (Emsam), Tranylcypromine (Parnate)] | Important drug interactions risking serotonin syndrome |
| Patients taking astemizole, cisapride, mesoridazine, ramelteon, or terfenadine | Contraindicated due to hepatic CYP3A4 interaction |
| Patients taking phenytoin or valproic acid | Potential interaction leading to seizure |
| Patients who are taking mirtazapine, melatonin, tramadol, or triptan medications | If these drugs could not be held, there was a risk of drug interaction increasing levels of these medicines |
| Participants taking alosetron, clozapine, flutamide, mexiletine, olanzapine, rasagiline, ropinirole, tacrine, theophylline, tizanidine, triamterene | Drugs are primarily metabolized by CYP1A2, which is inhibited by fluvoxamine. |
| Diazepam or alprazolam users | Due to interactions, we recommend reducing the dose by 25% unless the patient has a known seizure disorder (in which case they were excluded). |
